# Platelet hyperactivity in patients of vascular dementia

**DOI:** 10.1101/2023.07.12.23291574

**Authors:** Priya Dev, Mohammad Ekhlak, Ashish Yadav, Debabrata Dash, Abhishek Pathak

## Abstract

**Background:** Platelet-monocyte (PMA) and platelet-neutrophil aggregations (PNA) are critical in causing acute inflammatory reactions favoring vascular dysfunction. However, the precise pathophysiological link between Platelet-leukocyte aggregates and Vascular Dementia (VaD) remains undetermined. Our study aimed to investigate whether platelet hyperresponsiveness is independently associated with a predictor of VaD.

**Methods:** Platelet from 19 VaD patients and 18 age-matched healthy controls were subjected to different investigations.

**Result:** PMA, PNA, P-selectin externalization, and intracellular free Ca^+2^ ([Ca^+2^_i_]) flux were evaluated either in whole blood or in platelet-rich plasma. The result revealed that PMA, PNA, P-selectin, and [Ca^+2^]_i_ were found to be significantly outnumbered in the VaD group (4.1, 2.8, 2.7, and 2.5 times higher) compared to the control group with p-value <0.001, <0.001, <0.001, and 0.001 at 95% CI = 31.164 to 54.855, 8.653 to 22.793, 35.064 to 94.369 and 8747.015 to 28829.618 respectively.

**Conclusion:** Patients with Vascular Dementia have increased platelet leucocyte interaction, and PMA has the most significant prediction of vascular dementia than in subjects of healthy control. Thus, platelets in VaD patients switch to a ‘hyperactive’ phenotype.

## INTRODUCTION

Vascular dementia (VaD) is a type of dementia marked by significant cognitive impairment caused by an ischemic or haemorrhagic stroke or hypoperfusion of brain areas involved in memory, cognition, and behaviour. The following lesions are the most common causes of vascular dementia: ischemic lesions affecting large or small vessels (such as single strategic strokes, multi-infarct dementia, small-vessel disease, lacunas, and venous occlusions); or hypoperfusion lesions (including global or selective ischemia, border-zone infarcts, and partial white matter ischemia).[1] VaD is the second most prevalent subtype of dementia after Alzheimer’s disease (AD), contributing to around 15% to 20% of dementia cases in North America and Europe, with slightly higher estimates of over 30% in Asia and developing nations.[2–4] The overall risk of dementia following stroke varies greatly, ranging from 7% in population-based studies of first-ever stroke in previously non-demented people to over 40% in hospital-based studies of recurrent stroke with pre-stroke dementia.[5]

Stroke is still a well-known cause of dementia, increasing the risk by nearly two-fold.[6,7] Stroke features, particularly severity and location, contribute to a significant portion of the variability in dementia risk following a stroke.[8–11] Depending on education and premorbid cognitive performance, pre-existing medial temporal lobe atrophy, family history of dementia, and comorbidities, patients may resist mental deterioration even after severe strokes.[5,8] Although dementia following a stroke is highest in the subacute period, it continues for several years afterward, implying either a role for cognitive reserve or standard etiological variables, such as cerebral small vessel disease. As previously stated, a closer examination of the brain in old age frequently reveals a variety of pathologies, ranging from vascular pathologies such as macroscopic infarcts, microbleeds, micro infarcts, and amyloid angiopathy to parenchymal amyloid and tau deposits, TDP (TAR DNA-binding protein)-43, hippocampal sclerosis, and - synucleinopathies.[12–14] With a typical age of onset of dementia in the community of over 80 years, nearly every patient coming to a non-specialized memory clinic is likely to have a multifactorial aetiology of cognitive loss.[15]

Progressive VaD often culminates in acute thrombotic events. Crosstalk between platelets and leukocytes influences primary and acute inflammatory reactions, favouring secondary thrombosis at embolization locations. Leukocytes influence the inflammatory response as well as vascular dysfunction. Platelet-leukocyte interaction contributes to the various phases of ischemic stroke. Increased circulating levels of platelet neutrophil aggregates (PNAs) and platelet monocyte aggregates (PMAs) are linked to direct cellular contacts between platelets and innate leukocytes during the acute phase of stroke.[16] PNAs rise in the first three days following an ischemic event, while PMAs are mostly observable on day two.[17] Interestingly, platelet sequestration requires neutrophil adherence, suggesting that platelet dysfunction is not the primary cause of post-ischemic vascular dysfunction.[18] The interaction between platelets and leukocytes is thought to promote inflammatory processes and tissue damage by increasing leukocyte activation and recruitment.[19] Platelet-leukocyte interactions are mediated by P-selectin (CD62P) and CD40L-CD40, which are elevated in platelets of patients with acute stroke or transient ischemic attack;[20] enhance brain damage by facilitating leukocyte activation recruitment, increasing BBB permeability, and increasing the infarct volume.[18,21]

The precise pathophysiological link between Platelet-leukocyte aggregates and VaD yet remains undetermined. Although platelet-leukocyte interactions have been studied in ADs patients, the study in VaD will help us in differential diagnosis with a more precise subtype diagnosis beyond a coarse clinical cognitive phenotype. This study aims to study the association of PMA, PNA, P-selectin, and calcium levels as a part of platelet function in vascular dementia patients compared to healthy control. This study will guide us towards a better approach in treatment decisions depending upon pathways implicated in VaD patients, notably those of vascular origin. So, we aim to study the crosstalk between platelet and leukocytes in VaD patients.

## METHODOLOGY

### Patient selection

Vascular dementia was diagnosed based on the National Institute of Neurological Disorders and Stroke-Association Internationale pour la recherche et l’Enseignement en Neurosciences (NINDS AIREN) Criteria; Cognitive functions were assessed by MMSE, and a complex cognitive function was evaluated. All consecutive patients with vascular dementia who were admitted or visited the outpatient department were enrolled in the study. General inclusion/exclusion criteria were as follows:

### Inclusion criteria

(1) Control subjects: these individuals were fully independent in the activities of daily living (ADL), and instrumental activities of daily living (IADLs); (2) Probable VaD: meet the National Institute of Neurological Disorders and Stroke-Association Internationale pour la Recherche et l’Enseignement en Neurosciences (NINDS AIREN) Criteria; (Diagnosis of type 2 diabetes (DM) was based on American Diabetes Association criteria such as fasting plasma glucose of 7.0mmol/L, current treatment with a hypoglycaemic agent, casual glucose of 11.1mmol/L. For the controls or the patients with impaired fasting glucose DM was diagnosed if a 2-hour post-glucose level after an approx. 75-g oral glucose tolerance test 11.1mmol/L. Hypertension was diagnosed if systolic blood pressure >140mmHg and Diastolic pressure >90mmHg, or as antihypertensive treatment. Two readings were taken with a 5-minute interval between measurements. The mean of the two readings was recorded. CAD (coronary athero-sclerotic heart disease) was defined as the occurrence of a non-fatal myocardial infarction, percutaneous coronary angioplasty, or other forms of acute or chronic ischemic heart disease.

The diagnostic evaluation included: Non-Contrast Computed Tomography (NCCT) or Brain Magnetic resonance imaging (MRI), hemogram, biochemical tests, electrocardiogram, transoesophageal echocardiography, vascular imaging (intra- and extra-cranial vessels), assessment of prothrombic state. The patient also underwent 24 h Holter monitoring to rule out any atrial fibrillation.

### Exclusion criteria

Exclusion criteria for this study included other psychiatric diseases, chronic alcoholism, tumour, infection, haematological disorders, atrial fibrillation, chronic liver and kidney diseases, abnormal vitamin B12 or thyroid function tests, depression (as indicated by a score of 410 on the Geriatric Depression Scale), Parkinson’s disease, therapeutic use of anticoagulant for various medical conditions and dementia treatment.

### Sample preparation

Blood from patients diagnosed with VaD (n=19) was collected within seven days of VaD onset from the forearm venepuncture with minimal stasis to avoid platelet stimulation. Sample from age-matched healthy control (n=18) with no history of a relevant disease or medication within a year was collected after obtaining written informed consent, strictly as per recommendations and approved by the Institutional Ethical Committee of the Institute of Medical Sciences, Banaras Hindu University. The study methodologies conformed to the standards set by the Declaration of Helsinki. Blood was processed within 20 min of collection. Whole blood was employed for the platelet-leucocyte interaction experiment. Blood was centrifuged at 100×*g* for 20 min at room temperature to obtain platelet-rich plasma (PRP), used for P-selectin externalization and intracellular calcium measurement studies.[22]

### Platelet monocyte and neutrophil aggregation

Fresh human blood (20 µl) was added to a cocktail containing 10 µl each from APC-anti-CD41a (platelet-specific) (Hu CD41a APC HIP8; Cat no.: 559777) and FITC-anti-CD14 (leukocyte-specific) antibodies (Hu CD14 FITC M5E2; Cat no.:555397) and mixed gently. RBCs were lysed with 800 µl FACS lysis solution (1X, BD Biosciences) (BD FACS Lysing Solution 10X 100ML IVD; Cat no.: 349202) for 10 min at RT. Leucocyte-platelet interaction was analysed on a flow cytometer (FACS Calibur, BD Biosciences) within 30 min. Side scatter voltage was set at 350 with a threshold of 52 V, and amorphous gates were drawn to encompass neutrophils and monocytes separate from noise. A dot plot of side scatters (SSC) versus log FITC-CD14 fluorescence was created in the CellQuest Pro software. Amorphous gates were drawn for monocyte (high fluorescence and low SSC) and neutrophil (low fluorescence and high SSC) populations. All fluorescence data were collected using 4-quadrant logarithmic amplification for 1000 events in either neutrophil or monocyte gate from each sample and analysed using CellQuest Pro Software.

### c α-granules

Secretion from platelet α-granules in response to a stimulus was quantified by surface expression of P-selectin (CD62P). Briefly, 50 µl PRP were taken, and cells were stained with FITC-labelled anti-CD62P antibody (Hu/NHP CD62P PE AC1.2; Cat no.: 550561) (5 % v/v) for 30 min at room temperature in the dark. Samples were suspended in sheath fluid and subjected to flow cytometry. The acquisition was performed on a constant number of particles (10,000 events), and the results were expressed as the mean of positive platelets.

### Measurement of intracellular free calcium ([Ca^+2^]_i_)

PRP (5 µl) was diluted in 495 µl buffer B (20 mM HEPES, 138 mM NaCl, 2.9 mM KCl, 1 mM MgCl_2_, 0.36 mM NaH_2_PO_4_, supplemented with 5 mM glucose, pH 7.4). Fluo-4/AM was added to each tube and was incubated for 30 min at room temperature. After appropriate gating of platelets using a flow cytometer (Becton Dickinson, model Accuri C6), events were analysed in the FL1 channel within the time-lapse of 1.5–4.0 min (in the upper right quadrant) following the addition of reagents. Baseline calcium levels were recorded for 60 s, followed by the addition of 1 mM CaCO_3_ and five µM TRAP using gel-loading pipette tips as described by Mallick et al., 2015.[23]

### Flow cytometry acquisition and interpretation of data

Flow cytometry was performed using a FACS Calibur instrument with CELLQuest software (Becton Dickinson Immunocytometry Systems). Forward and side scatter measurements were made with gain settings in linear mode to analyse platelet-monocyte or platelet-neutrophil interactions. The monocyte and the neutrophil populations were thus easily distinguished (Figure 1A, 1B). A minimum of 2000 monocytes and 5000 neutrophils were acquired for each determination. A three-color analysis was used for the simultaneous detection of platelet-leukocyte aggregates and platelet-leukocyte activation. Monocytes and neutrophils were further gated into a CD41a-positive platelet-bound population, gate two, and a CD41a-negative platelet-free population, gate three (FL1 versus FL4, Figure 1A, 1B). Unbound platelets were measured with forwarding and side scatter signals in logarithmic mode. Ten thousand events were acquired with a live gate on FSC versus SSC. Results were compared to antibody-matched controls and expressed as positive percentage cells or mean fluorescence intensity (MFI) after subtracting the control antibodies.

**Figure 1:**
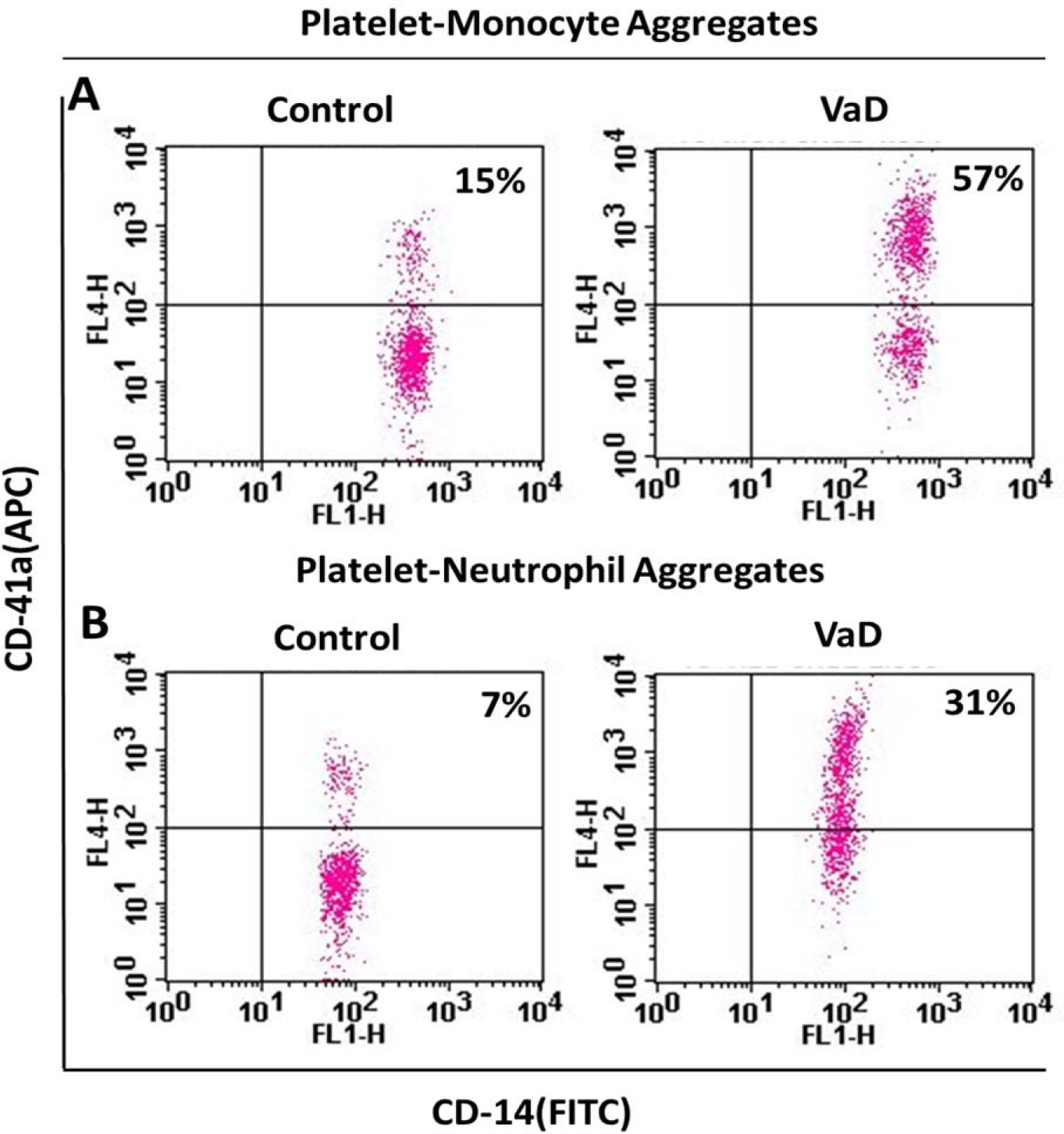
Detection of platelet-monocyte and platelet-neutrophil aggregates in circulation. A and B, flow cytometric analysis of monocyte-platelet and neutrophil-platelet aggregates, respectively, in whole blood stained with platelet-specific marker, anti-CD41a-APC and neutrophil/monocyte-specific marker, anti-CD14-FITC. Samples were obtained from control and different vascular dementia (VaD) groups as indicated. Amorphous gates were drawn for monocyte (high fluorescence and low SSC) and neutrophil (low fluorescence and high SSC) populations. Numbers in right upper quadrant represent percent of aggregates. C and D, corresponding scatter plot bar diagrams representing mean percent of PMA and PNA, respectively.

### Ethical Statement

The Institutional ethical committee of the Institute of Medical Sciences, Banaras Hindu University, UP, India, received approval for a study protocol. All participants, before enrolment, read the study protocol and signed informed consent to have been included in the study. The study was conducted by the Declaration of Institutional ethics committee.

### Statistical Evaluation

The statistical analysis was performed with the SPSS v16. The obtained data were expressed as mean and standard deviation. The student’s t-test was performed to determine statistical significance. The level of p < 0.05 was considered the threshold for statistical significance. ROC curve was drawn in a post hoc analysis to determine the cut-off value of the platelet-leukocyte experiment variables for the diseased and control group. Further, principal component analysis (PCA) to select the explanatory variable for the logistic regression model.

## Results

The study involved 19 VaD patients (with a mean age of 63.63±12.35 years) and 18 age and gender-matched healthy control subjects (with a mean age of 60.22±14.17 years). Their demographic data is represented in table 1.

**Table 1:** Baseline demographic and clinical data of VaD patients and Control.

| Sl. No. | Variables | VaD (N=19) | Control (N=18)<br>(Mean±SD) | P-Value | 95% CI |  |
| --- | --- | --- | --- | --- | --- | --- |
|  |  |  |  |  | Lower | Upper |
| 1 | Age (mean ± SD) | 63.63±12.35 | 60.22 14.17 | 0.4400 | -5.447 | 12.266 |
| 2 | NIHSS | 16.42±5.83 |  |  |  |  |
| 3 | GCS (Out of 15) | 12± 2.91 |  |  |  |  |
| 4 | PMA (%) | 57.5 | 14.1 | 0.0000 | 31.164 | 54.855 |
| 5 | PNA (%) | 24.5 | 8.8 | 0.0001 | 8.653 | 22.793 |
| 6 | P-sel (MFI) | 103.140± 54.971 | 38.423± 29.334 | 0.0001 | 35.064 | 94.369 |
| 7 | Ca<sup>++</sup> | 30948.595±<br>20014.593 | 12160.278±436.038 | 0.0006 | 8747.015 | 28829.618 |
| 8 | Male (%) | 15(78.9) | 16(88.9) | 0.412 |  |  |
| 9 | History of Stroke (%) | 19(100) |  |  |  |  |
| 10 | Location of the stroke (%) |  |  |  |  |  |
|  | PACI | 17(89.5) |  |  |  |  |
|  | LACI | 2(10.52) |  |  |  |  |
| 11 | TOAST classification (%) |  |  |  |  |  |
|  | SVD | 10(52.6) |  |  |  |  |
|  | Cardioembolic | 7(36.8) |  |  |  |  |
|  | Cryptogenic | 2(10.5) |  |  |  |  |
| 12 | History of comorbidities (%) |  |  |  |  |  |
|  | HTN (Yes) | 10(52.6) | 0 | 0.000 |  |  |
|  | DM(Yes) | 2(10.5) | 0 | 0.213 |  |  |
|  | Dyslipidemia | 1 | 0 | 0.324 |  |  |
| 13 | Fazekas (%) |  |  |  |  |  |
|  | 0 |  |  |  |  |  |
|  | 1 | 5(26.3) |  |  |  |  |
|  | 2 | 9(47.4) |  |  |  |  |
|  | 3 | 5(26.3) |  |  |  |  |
| 14 | Hemisphere (%) |  |  |  |  |  |
|  | Right | 2(10.6) |  |  |  |  |
|  | Left | 3(15.8) |  |  |  |  |
|  | Bilateral | 14(73.7) |  |  |  |  |
| 15 | MRS (%) |  |  |  |  |  |
|  | 3 | 2(10.5) |  |  |  |  |
|  | 4 | 5(26.3) |  |  |  |  |
|  | 5 | 11(57.9) |  |  |  |  |
|  | 6 | 1(0.3) |  |  |  |  |
| 16 | MRS Follow-up (%) |  |  |  |  |  |
|  | 2 | 1(5.3) |  |  |  |  |
|  | 3 | 4(21.1) |  |  |  |  |
|  | 4 | 5(26.3) |  |  |  |  |
|  | 5 | 3(15.8) |  |  |  |  |
|  | 6 | 6(31.6) |  |  |  |  |

Out of 19 VaD patients, 79% were male; all had a history of ischemic stroke; 17 patients had partial anterior circulation infarct (PACI) and two patients had Lacunar infarct (LACI); 10 patients were Small vessel disease (SVD), seven were cardioembolic, and 2 were cryptogenic stroke in origin in TOAST classification; 52.6% had a history of hypertension, and 26.3% patients had periventricular white matter (PWM) changes with Fazekas grade 3. (Table 1)

An independent sample t-test was used to analyse the PMA, PNA, P-selectin, and [Ca^+2^]_i_ level between VaD patients and control subjects. The result revealed that PMA (fig 1A), PNA (fig 1B), P-selectin (fig 2A), and [Ca^+2^]_i_ (fig 2B) were found to be significantly higher in the VaD group (4.1, 2.8, 2.7, and 2.5 times higher) compared to the control group with p-value <0.001, <0.001, <0.001, and 0.001 at 95% CI = 31.164 to 54.855, 8.653 to 22.793, 35.064 to 94.369 and 8747.015 to 28829.618 respectively. (Table 1) (Fig 1 and 2).

**Figure 2.**
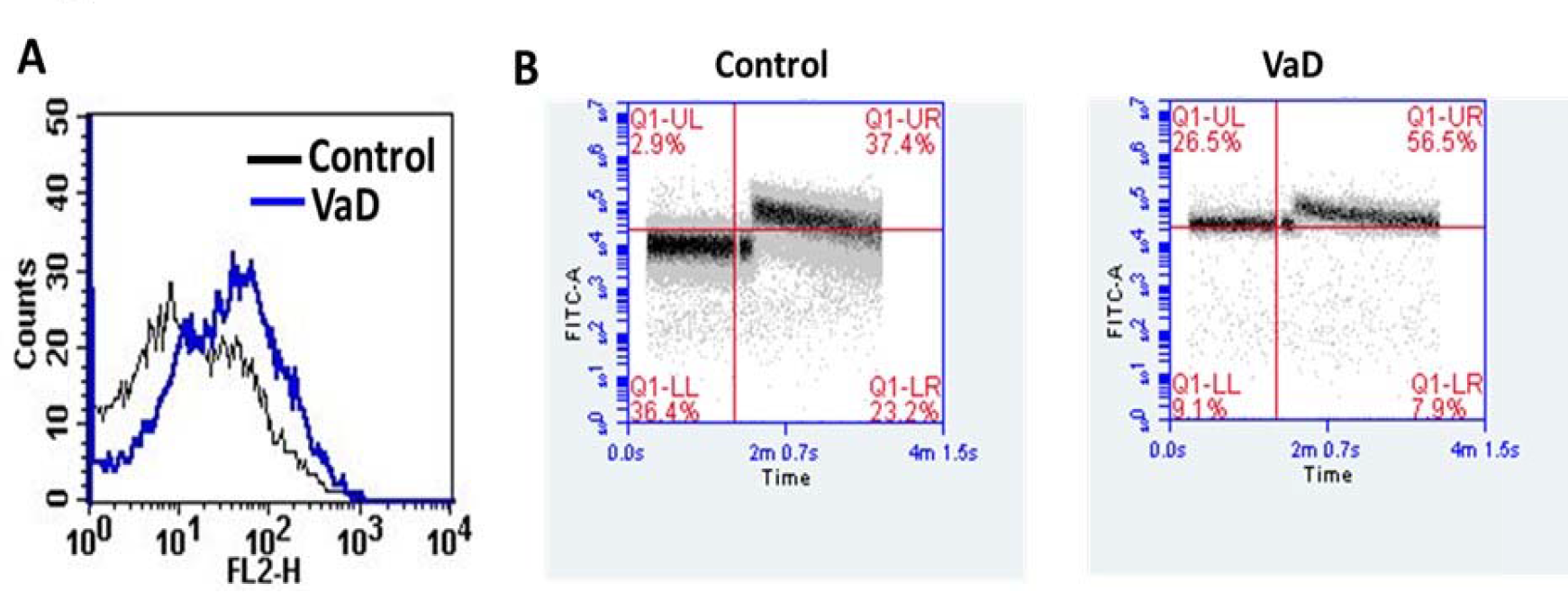
Exposure of P-selectin on surface of unstimulated platelets and determination of platelet population with raised intracellular calcium. A, flow cytometric analysis of binding of anti-P-selectin antibody to surface of unstimulated platelets from VaD group and control group as indicated (representing mean fluorescence intensity). B, flow cytometry of Fluo-4-labeled platelets representing cell population with high fluorescence intensity in control (n=18), and VaD (n=19) groups, respectively, (representing mean fluorescence intensity over 1.5 min of a population before TRAP addition).

The post hoc analysis was used to draw a ROC curve to determine the cut-off value for PMA (with the area under the ROC curve (AUC)= 0.988), PNA (AUC=0.944), p-selectin (AUC=0.819) and [Ca^+2^]_i_ (AUC=0.871) variables for discrimination VaD from the control group. Thus, the cut-off value was determined as 22.35% (sensitivity=94.7% and specificity=100%), 17.00% (sensitivity=78.9% and specificity=100%), 61.37 MFI (sensitivity=78.9% and specificity=78%), and 18778.5 MFI (sensitivity=78.9% and specificity=78%) for PMA, PNA, P-selectin and [Ca^+2^]_i_ variables respectively. (Table 2).

**Table 2:** Area under the curve (ROC table)

| Variables | Area | Standard-Error | P-Value | 95% CI |  |
| --- | --- | --- | --- | --- | --- |
|  |  |  |  | UPPER | LOWER |
| PMA | 0.988 | 0.013 | 0.000 | 0.962 | 1.015 |
| PNA | 0.944 | 0.034 | 0.000 | 0.878 | 1.011 |
| P-Selectin | 0.819 | 0.071 | 0.001 | 0.680 | 0.958 |
| Ca<sup>++</sup> | 0.817 | 0.061 | 0.000 | 0.752 | 0.990 |

The predictor variables PMA, PNA, P-selectin, and [Ca^+2^]_i_ were highly correlated the value of the Pearson correlation coefficient lies in the range from 0.65 to 0.84. The problem of collinearity was addressed using a multivariate approach called principal component analysis (PCA). PCA results suggest that the first component, i.e., PMA had the highest eigenvalues and then is PNA’s value. But in supplementary table 1, we can observe that in the control group there is a significant (P=0.0014) association (r=0.69) between PMA and PNA, and rest of the covariates are not correlated. One can observe a circular representation of a scatter plot from Supplementary Figure 1 representing a week correlation. Similarly, we can observe that in the diseased group, except for P-selectin and PNA rest, all the covariates are significantly associated with each other (Supplementary Table 2). The exact inference can be observed in Supplementary Figure 2, where a positive linear trend is observed between the covariates.

Thus, finally, a logistic regression model with the group as outcome variables (PNA, P-selectin, and [Ca^+2^]_i_), as an explanatory variable was developed however, the models suggest that out of them, only PNA as an explanatory variable was statistically significant. (Table 3) Table 3 shows that PNA is a significant predictor variable in explaining VaD disease and alone can explain 38% of the variation.

**Table 3:** Factor correlation and Logistic Regression analysis

| Variables | Factor correlation | Logistic Regression |  |  |
| --- | --- | --- | --- | --- |
| | | β-coefficient | 95% CI | |
|  | Eigen's value |  | UPPER | LOWER |
| PMA | 0.988 |  |  |  |
| PNA | 0.944 | 0.3822 | 0.028 | 0.042 |
| P-Selectin | 0.819 | -0.0022 | 0.913 | -0.042 |
| Ca<sup>++</sup> | 0.817 | 0.0002 | 0.093 | -0.000 |

## Discussion

Our result shows that all the platelet functions, namely, PMA, PNA, p-selectin, and [Ca^+2^] I, are significantly increased in VaD compared to control. The post-hoc analysis concluded that PMA had the eigenvalue in Vascular Dementia compared to the control. This is the first study with the above-mentioned parameters looking into the platelet activity in Vascular dementia vis-a-vis age and sex-matched control patients. The increased platelet activity has been implicated in the development of Alzheimer’s Disease but their role in the development of Vascular Dementia has not been studied adequately. Vascular Dementia has been implicated to have an underlying inflammatory cascade. The role of platelets during the inflammatory state interacts with leucocytes via various soluble mediators and direct cell-based interactions. Platelet has the highest affinity for macrophages and monocytes, followed by neutrophils. Cell-based interaction includes activation of CXCL4, which prevents monocyte apoptosis and, along with other cell-based receptors, namely CD40L and CD62P, promotes the production of cytokines and ROS. Increased platelet activity has a significant contribution to the development of vascular dementia. In a similar study done by Konstantinos Stellos et al. on 37 patients found that higher platelet activity was seen in patients with Vascular cognitive impairment similar to our study. Leucoariosis has independently been associated with the development of Vascular Cognitive impairment. Fujita et al. studied the platelet hyperaggregability in the development of leukoaraiosis in 73 patients and they concluded that patients with leukoaraiosis had significantly higher platelet hyperaggregability than patients without leukoaraiosis. In a similar study done by Nagamo Kuriyama et al., in 143 subjects with deep white matter lesions(DWML) with cognitive decline, found significant increases in hypertension and CD62P levels with increasing DWML grade. This study also observed a significant negative correlation between CD62P levels and MMSE scores. Our study did not directly look into DWML but our patient group also had patients with DWML and vascular cognitive impairment.

The increased platelet activity contributes to cognitive impairment through several mechanisms. Firstly, platelet-bound GPIIb-IIIa and P-selectin interaction lead to increased adhesion, rolling, recruitment, and diapedesis of platelets on endothelial cells, ultimately leading to vascular and perivascular inflammation leading to dementia progression. Secondly, it also leads to progressive carotid atherosclerosis, an independent risk factor for vascular dementia independently, via various secondary mechanisms, including vasoconstriction and hypoxia.

Our study had a few advantages; first, this was one of the first studies to look into the platelet leucocyte interaction in patients with Vascular dementia. Secondly, the assessor of platelet leucocytes was blinded to the samples of Vascular dementia and healthy control. Thirdly the patient was purely of Vascular dementia, and none of the patients with mixed dementia were included.

Our present study had a few major limitations. Firstly, the sample size of the patients was very small. Secondly, the other types of dementia could also have been chosen as a control, namely FTD, AD, and DLBD. Thirdly whether increased platelet interaction is the cause or the effect cannot be proven by the present study. To prove its causality, a prospective study with intermittent platelet activity with a larger population of patients with vascular risk factors developing into Vascular dementia should be studied.

However, this study indicates that drugs that would inhibit platelet leucocyte interaction may have the potential for therapeutic importance in patients with vascular dementia.

### Conclusion

Patients with Vascular Dementia have increased platelet leucocyte interaction, and PMA has the most significant prediction of vascular dementia than in subjects of healthy control. Inhibiting this platelet leucocyte interaction may have a therapeutic advantage. However, larger prospective studies with a larger population of vascular dementia are required to prove the present hypothesis.

## Data Availability

All data generated or analysed during this study are included in this article and its supplementary material file.

## Acknowledgements

I would like to thank BHU and Sir Sunder Lal Hospital for their infrastructural support.

## Statement of Ethics

**-Study approval statement:** This study protocol was reviewed and approved by Institutional Ethics Committee, Institute of Medical Sciences, Banaras Hindu University, Varanasi, India, and Reference number [Dean/2020/EC/2155]

**-Consent to participate statement:** WRITTEN informed consent was obtained from participants or legal guardian to participate in the study.

## Authors agreement

All authors have viewed and agreed to the submission

## Consent for publication

Not applicable

## Consent for publication

Not applicable

## Author Contributions

PD: Experiments, paper writing and editing; ME: experiments; AY: Statistical analysis; DD: provided instruments facility for Experiments; AP: conceptualization, paper editing, writing

---

**Supplementary Figure 1:**
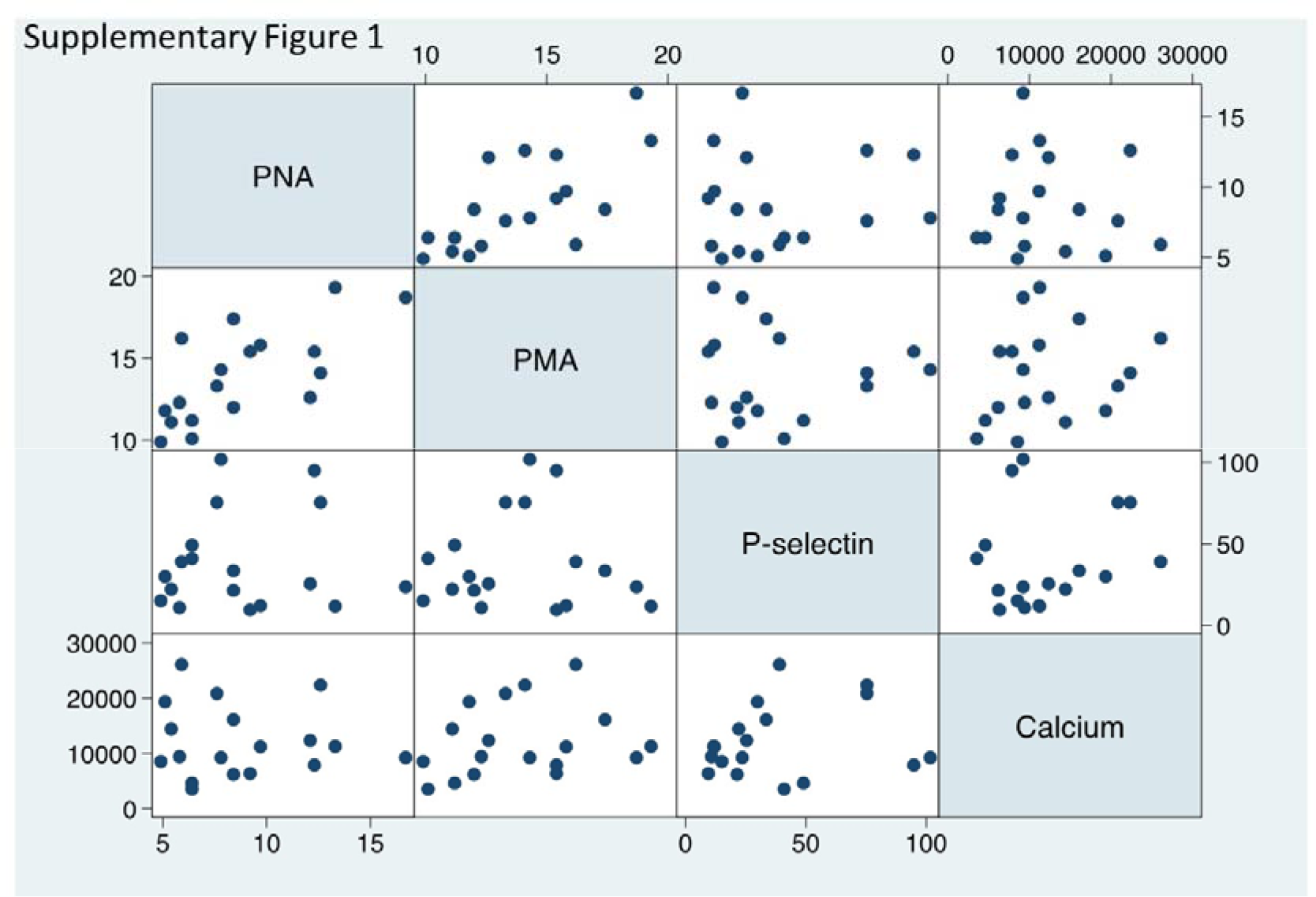
Scattered plot showing correlation between predictor variables in control group

**Supplementary Figure 2:**
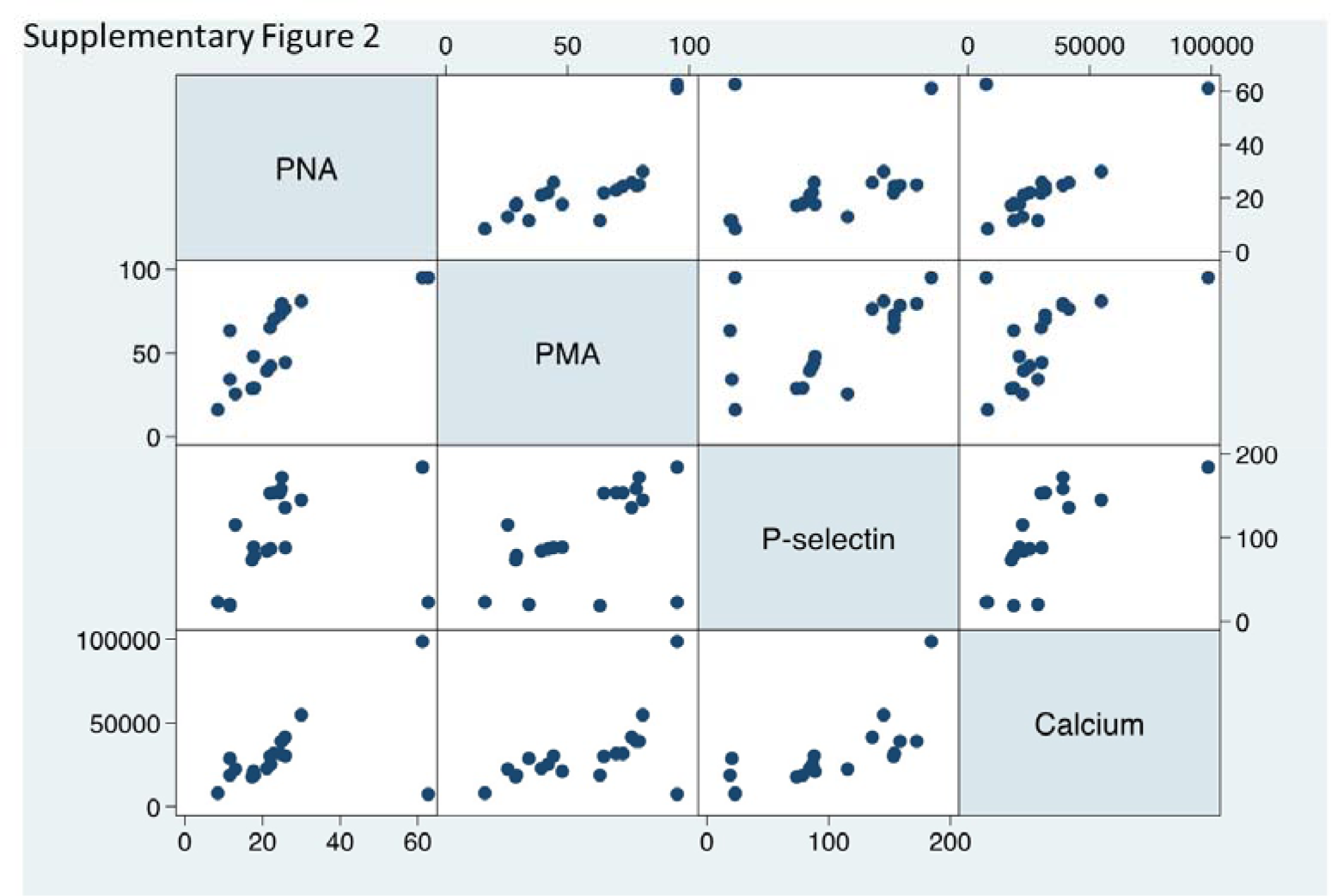
Scattered plot showing predictor variable in diseased group

**Supplementary Table 1:** Correlation between Predictors for Control group.

|  | PNA | PMA | P-selectin | [Ca<sup>+2</sup>]<sub>i</sub> |
| --- | --- | --- | --- | --- |
| PNA | 1.0000 |  |  |  |
| PMA<br>(p-value) | 0.6936<br>**(0.0014)** | 1.0000 |  |  |
| P-selectin<br>(p-value) | 0.0896<br>(0.7237) | -0.0124<br>(0.9610) | 1.0000 |  |
| [Ca<sup>+2</sup>]<sub>i</sub><br>(p-value) | -0.0708<br>(0.7802) | 0.2172<br>(0.3866) | 0.1808<br>(0.4727) | 1.0000 |

**Supplementary Table 2:** Correlation between Predictors for Case group.

|  | PNA | PMA | P-selectin | [Ca<sup>+2</sup>]<sub>i</sub> |
| --- | --- | --- | --- | --- |
| PNA | 1.0000 |  |  |  |
| PMA<br>(p-value) | 0.7447<br>**(0.0003)** | 1.0000 |  |  |
| P-selectin<br>(p-value) | 0.2724<br>(0.2592) | 0.5214<br>**(0.0192)** | 1.0000 |  |
| [Ca<sup>+2</sup>]<sub>i</sub><br>(p-value) | 0.5154<br>**(0.0239)** | 0.5689<br>**(0.0110)** | 0.6952<br>**(0.0010)** | 1.0000 |

